# Priorities setting in mental health research: a scoping review

**DOI:** 10.1101/2022.04.04.22273381

**Authors:** C. Benito-Amat, E. Güell, J. Molas-Gallart

## Abstract

**Objective:** Research processes are opening to stakeholders beyond the scientific community. We analyse the user involvement in the definition of research priorities in the field of mental health. Mental disorders represent a significant disease burden at a global scale and their identification and treatment involves caregivers, patients and related social groups such as family and friends. Therefore it is an area conducive to the application of participatory methods in priority setting. We present a scoping review of participatory methods in mental health priority setting for the period 2010-2020 to shed light on their spread and characteristics, the types of groups involved and the link with the priorities identified.

**Methods:** First we describe the eligibility criteria for the scoping review. We selected peer-reviewed documents published between 2010 and 2020 using MEDLINE/PubMed, PsycINFO, the Core Collection of the Web of Science and Scopus, applying controlled terms of search. We initially identified 330 documents from which we selected seventy-four after further discarding studies that were not specifically addressing priority setting in mental disorders research. We noted and classified the interest groups participating in every study.

**Results:** Priority setting partnerships are becoming the most frequent participatory instruments for priority setting in mental health. We identify regional differences in the extent to which such methods are being applied. When research beneficiaries participate in priority setting, prioritised research focuses on therapy, standards, education and psychology of mental disorders. When participation is limited to scientists, therapy, diagnosis, methods and standards, receive more attention.

## Introduction

There is a growing trend to open research and engage citizens into research processes. Citizen involvement in part or in the entire course of scientific inquiries has been described through concepts like “participatory research”, “research partnerships”[1], “integrated knowledge translation”[2], “participatory action research”[3], and “community academic partnership”[4] among others.

Despite some criticism[5,6], participatory approaches in health research have been found to improve the value of health research by shaping and informing the purpose and scope of research with a deeper understanding of user needs[7]. The participation of patients and other stakeholders can enrich not only the interpretation and translation of research results, but can also lead to research results better tailored to user^3^ needs[8]. In addition, co-governance with users opens information flows about barriers and resources needed for health research[9]. Similar conclusions are reported by the systematic review by Brett et al. (2014)[10]. Accordingly, a significant number of initiatives promoting participatory approaches have emerged like, for instance, the INVOLVE Program of the UK National Institute of Health Research (https://www.invo.org.uk/) established in 1996, the James Lind Alliance Priority Setting Partnerships[11] set up in 2004, the Patient-Centered Outcomes Research Institute, established in 2010[12], the Canada’s Strategy for Patient-Oriented Research launched in 2011, and the International Collaboration for Participatory Health Research, active since 2013[9].

Our focus will be on the participation in research priority setting in the field of mental health. The degree of involvement in research processes ranges from non-participation, to symbolic participation and engaged participation[13]. In the higher level of engagement (“engaged participation”), patients, caregivers and other interest groups dictate the priority setting for research choices, influence the design of the project activities and participate in the interpretation of research findings and their implications. Thus, the participation of social groups and communities provides a different way of identifying research gaps, determining priorities in health research[14], and reducing research initiatives not adjusted to users needs[15]. This social participation is particularly relevant in the field of mental disorders, which represent a significant burden of disease at a global scale[16]. Mental disorders are socially identified, have social antecedents or causes, and they have comprehensive social consequences[17]. Further, there is a significant gap between the problems posed by current mental disorders needs, and the resources and knowledge available to tackle them. According to the WHO [18], mental disorders account for 13% of the total global burden of disease and are projected to become the leading cause of mortality and morbidity by 2030. Therefore, there is a growing pressure to increase the promotion of mental health and the prevention of mental disorders, while their social aspects suggest a special rationale for the use of participatory methods in research priority setting.

Have participatory approaches been applied to priority setting in mental health? If so, which participation methods and approaches have been used? What type of priorities do they identify? Who are the most frequent stakeholders, consumers or groups consulted? Does the participation of different groups make a difference? We provide an answer to these questions through a scoping review of recent articles, following the PRISMA adaptation proposed by Tricco et al. (2018)[19].

## Method

Following the checklist proposed by Tricco et al. (2018), we first describe the documents’ eligibility criteria, then, we select the bibliographic sources and determine the search strategy used. Finally, we identify the variables extracted from each selected document.

### Eligibility criteria

We selected peer-reviewed documents published between 2010-2020. Peer-reviewed journal papers were included if either a) they considered different options and methods for setting priorities in mental disorders research, or b) they defined research priorities. Meeting abstracts were excluded from the results, although proceedings papers, later published as journal articles, were considered eligible.

### Information sources and search

In March 2020 we searched the following bibliographic databases: MEDLINE/ PubMed, PsycINFO, and the Core Collection of the Web of Science and Scopus. We adjusted search strategies for every database, taking into account that specialised databases have a controlled vocabulary. We then drew a combination of descriptors from the controlled vocabularies and from terms in articles’ titles. Further, journal subjects of classification were taken as the research context and combined with terms from the papers titles. Searches were performed by a member of the team long experienced in information retrieval from bibliographic databases.

First, we used the category “Mental disorders” in MEDLINE/PubMed)(F03 in the Medical Subject Heading (MeSH) tree), with more than 200 specific terms. As a specialised database, for PsycINFO we used the title term “research priorit*”. In addition, the natural language terms used in the previous MEDLINE search were combined with the entry “3200: Psychological & Physical Disorders” of the PsycINFO Classification Code System. Articles were retrieved through Web of Science from journals classified under the subject categories *Psychiatry* and *Substance abuse*. The average annual number of papers published by the combined set of about 170 journals in the period covered by our study (2010-2020) is 18,000. Subject categories were combined with the title terms list. Finally, Scopus had no subject categories specific enough for setting a context for mental disorders research. Therefore, we added the title terms used in the MEDLINE search with journal titles bearing the terms “addict*”, “Psychiatry” or “Mental”.

Second, among its term variants, we selected the following: “research priorities”, “research agenda”, “research portfolio”, “research framework”, “participation in research” or “stakeholders”. After removing duplicate entries, we selected 330 documents published from 2010 onwards.

### Selection of sources of evidence

A two-round selection process was then used to select publications from this initial set of documents. In the first round, titles and abstracts were examined by two members of the team, who judged their relevance. A first group of articles clearly addressed the topic of setting priorities on mental disorders research and were consequently selected. We rejected a second group of articles either because they were 1) unrelated to research issues; 2) mainly focused on training and capacity building of research personnel; 3) commentaries on other papers or just giving subjective opinions; 4) dealing with service provision, or 5) announcing future publications on the subject. Finally, we selected for further examination a third group of articles whose abstracts did not provide enough information. In a second round, we examined the full text of all selected studies as well as those with ambiguous abstracts, adding new relevant publications to the selected group. Inconsistencies between reviewers detected after the first round were solved through this second round.

A total of seventy-four sources of evidence were finally selected.

### Data charting process

We downloaded the study references in a bibliographic management system (Zotero), and we imported them into relational tables to process and chart the data. One reviewer charted the data and submitted the tables to a second reviewer, who proposed additions or amendments.

We used an abridged version of the reporting guideline for priority setting of health research (REPRISE) proposed by Tong et al. (2019)[20] to identify useful variables in the selected documents. With regard to the context and scope of the studies, we analysed the geographical scope, the health area and focus, the intended beneficiaries, and the research area. In addition, we processed the framework, methods or protocols used for collecting initial priorities and charted the priorities list proposed by the stakeholders.

### Data items

Since not all articles mention their geographical scope, we derived it from the author’s affiliation data, noting each author’s country or countries. To determine the articles’ focus, we used the controlled set of terms offered by the US National Library of Medicine (NLM), the Medical Subject Headings (MeSH), and its hierarchical structure to classify the conditions into groups.

We characterized the research beneficiaries according to the age, gender and ethnic group of the individuals targeted by the study. The methods used to elucidate stakeholders’ views were also registered along with the protocols or frameworks employed.

We noted and classified the interest groups participating in every study. Finally, we recorded the research priorities identified in every paper grouping them also under the MeSH subheadings that MEDLINE applies to the mental disorders literature. We extended the definition of the subheading “Psychology” to include references to the psycho-social aspects of the diseases. Several examples of this procedure are given in Table 1.

**Table 1.**
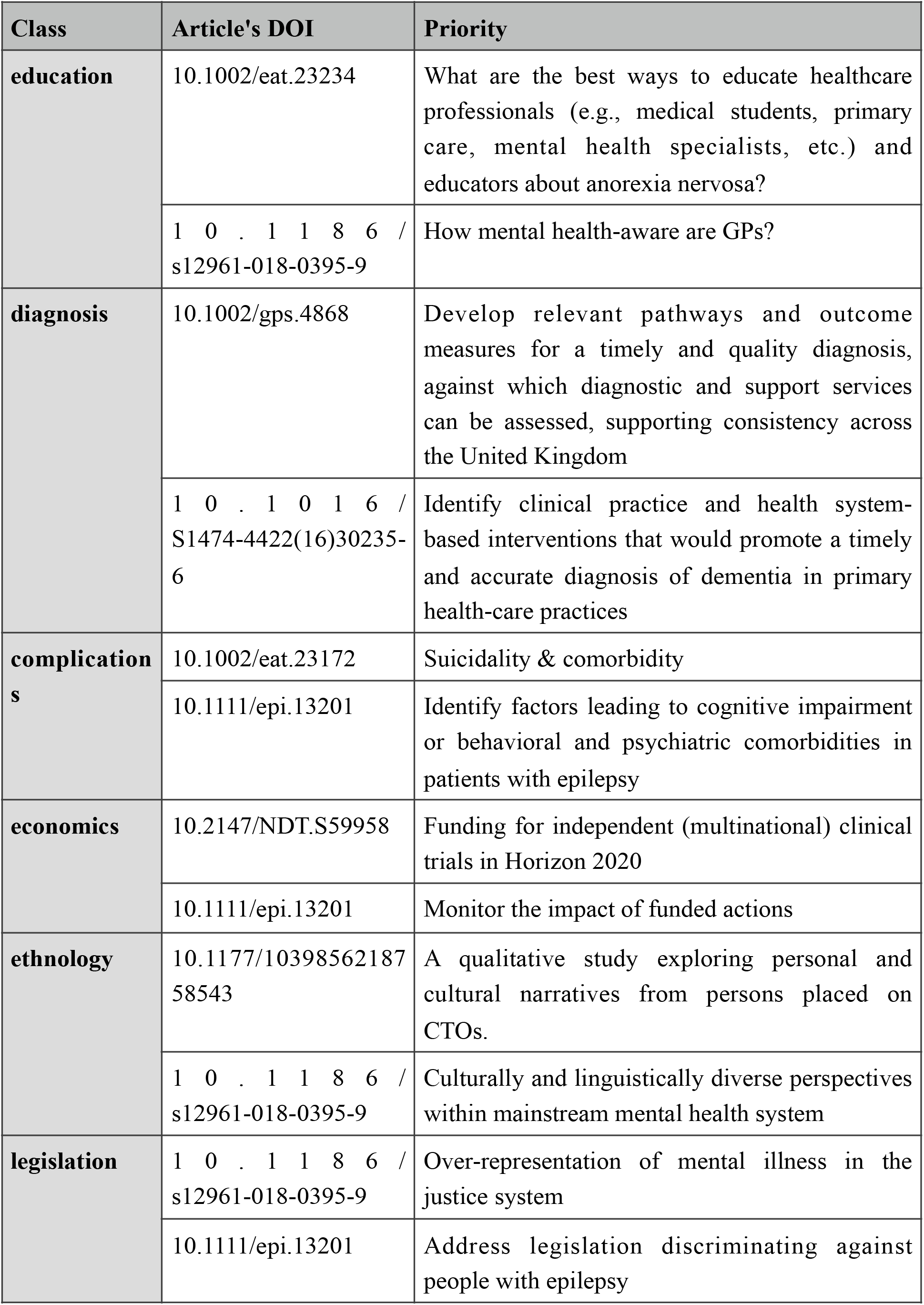
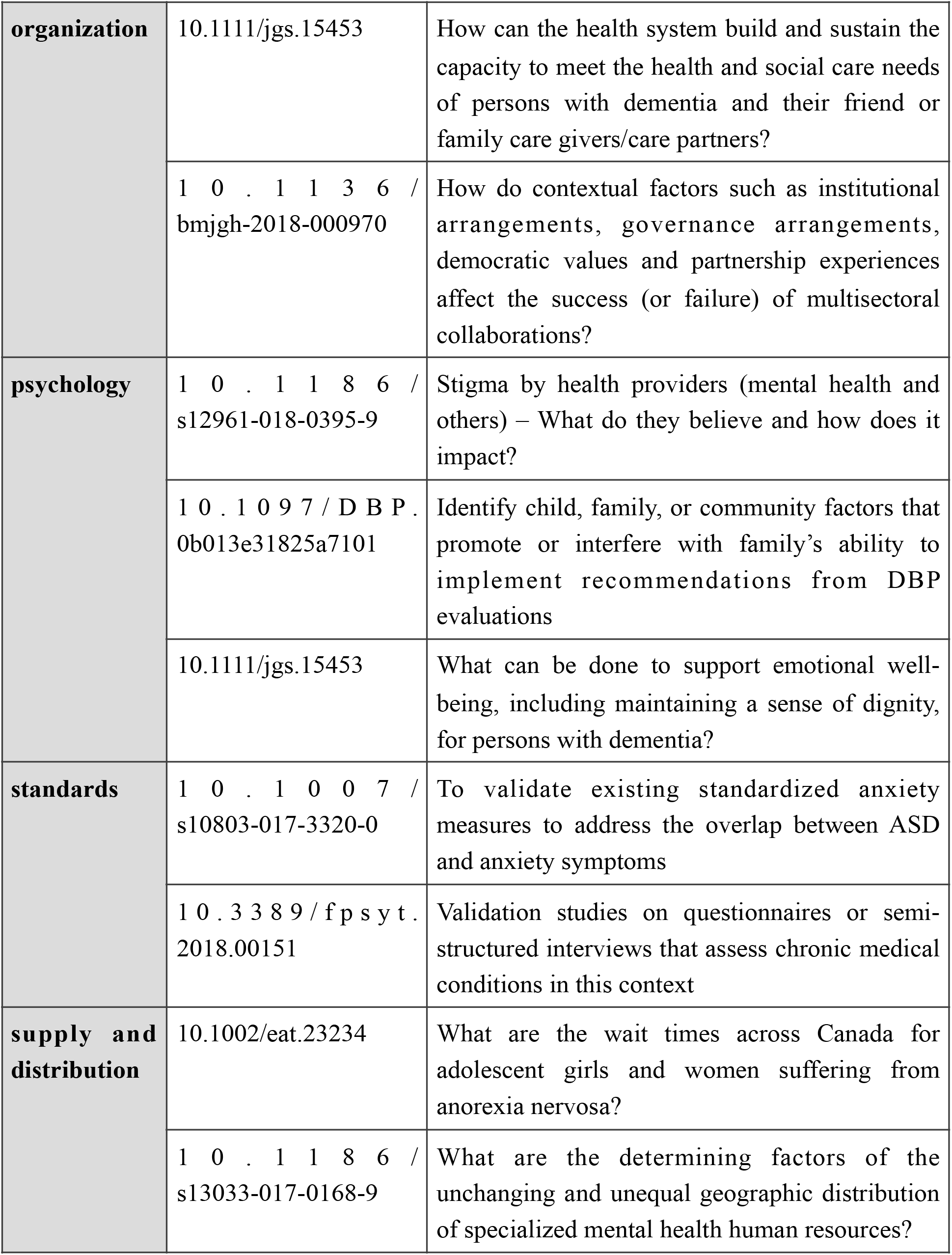
Some examples of priorities translated into classes.

## Results and discussion

The seventy-four selected articles are listed in the Supplementary Material. They can be classified into two main groups: (1) fifty-five original papers whose results are lists of research priorities; and (2) nineteen non-original articles that either review other studies or apply an authoritative method to recommend adopting priorities in the research of a given condition. This distribution fits the three approaches devised by Kelber et al. (2019)[14] as methods to identify research gaps/priorities: the systematic review of literature, the resource to authoritative sources, and the participation of social groups and communities. Our analysis focuses on the latter group, although we also describe the procedures followed in the former two.

### Geographical scope of the studies

The fifty-five original studies have been contributed by authors from forty-two different countries (see map in Figure 1). The United Kingdom leads with twenty contributed papers, followed by USA (16) Australia (15) and the Netherlands (11). Two thirds of the fifty-five studies are produced by authors from a single country. These are mainly consensus building papers, produced through research consortia or collaborative projects. There are, however, some important exceptions. Eighteen countries are listed in the affiliation data of the article by Khandelwal and cols. (2010)[21], a consensus building paper related to the Global Network for Research in Mental and Neurological Health. The ROAMER (Road Map for Mental Health Research in Europe) provides the background for Forsman (2015)[22], and Wykes (2015)[23] contributed by collaborators from fourteen and nine different countries, respectively. The Mental Health and Psychosocial Support in Humanitarian Settings[24], the European Forum for Epilepsy Research[25], and the European Association of Psychosomatic Medicine[26] are supranational efforts.

**Figure 1.**
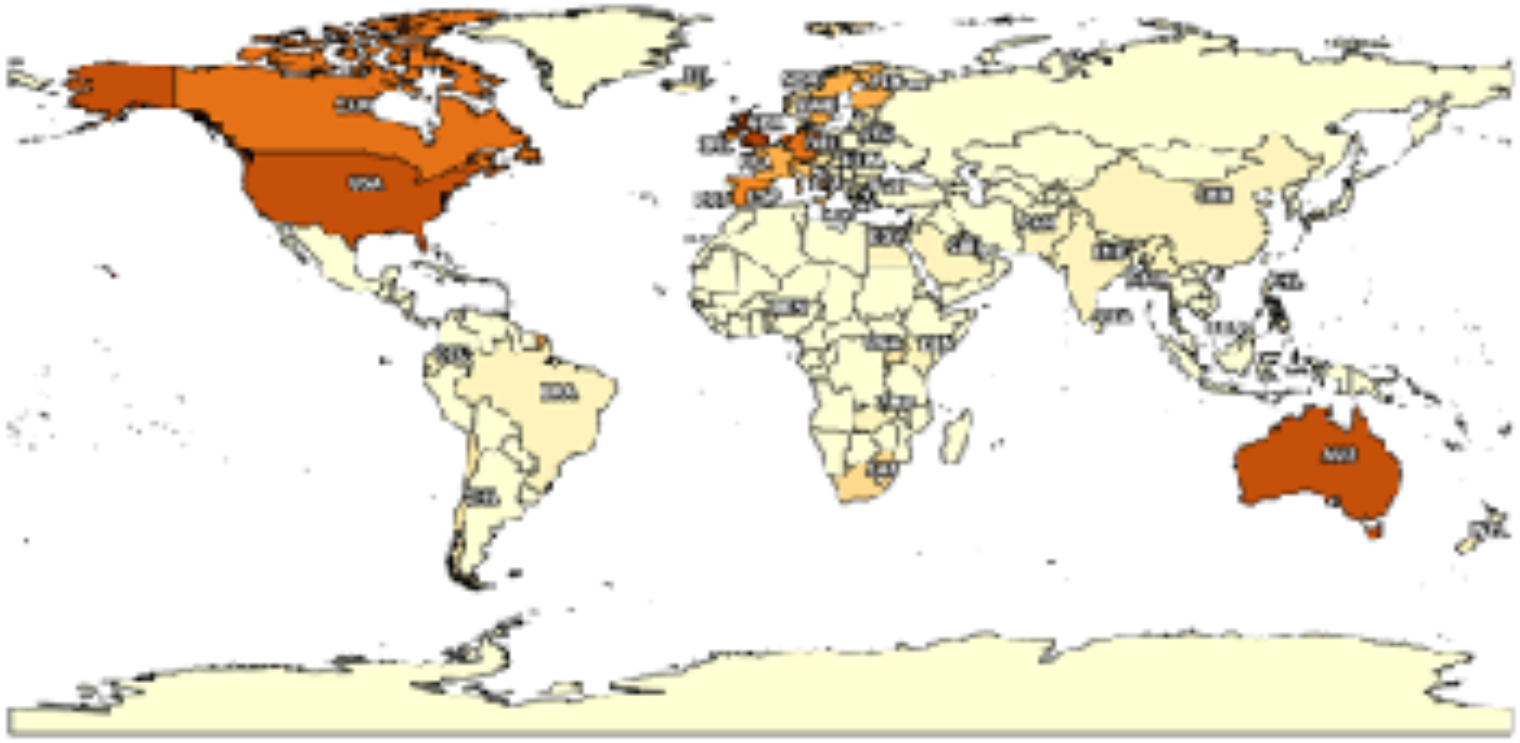
Geographical distribution of the studies

### Health area and focus

Forty studies set research priorities on twenty-seven specific disorders. (see Table 2 and the online Supplementary Material).

**Table 2.** Mental disorders studied

| Disorders | Studies |
| --- | --- |
| **Alzheimer Disease** | Gove, Dianne (2018); Liggins, Charlene (2014) |
| **Anorexia Nervosa** | Obeid, Nicole (2020) |
| **Attention Deficit Disorder with Hyperactivity** | Gaynes, Bradley N. (2014); Jacobson, Stella (2016) |
| **Autistic Disorder** | Clark, Megan (2020); Frazier, Thomas W. (2018); Pellicano, Elizabeth (2014); Russell, Ginny (2018); Shattuck, Paul T. (2018); Tomlinson, Mark (2014); Vasa, Roma A. (2018) |
| **Bipolar Disorder** | Banfield, Michelle A. (2011); Banfield, Michelle A. (2014); Maassen, Eva F. (2018) |
| **Brain Injuries, Traumatic** | Clavisi, Ornella (2013) |
| **Delirium** | Pandharipande, Pratik (2017) |
| **Dementia** | Bethell, Jennifer (2018); Iliffe, Steve (2013); Kelly, Sarah (2015); Law, Emma (2013); Leroi, Iracema (2019); Miah, Jahanara (2019); Pickett, James (2018); Schneider, Lon S. (2016); Shah, Hiral (2016) |
| **Depressive Disorder** | Banfield, Michelle A. (2011); Banfield, Michelle A. (2014); Topooco, Naira (2017) |
| **Developmental Disabilities** | Blum, Nathan J. (2012); Camden, Chantal (2019); Kramer, Jessica M. (2019) |
| **Dyssomnias** | Bassetti, C. L. (2015) |
| **Epilepsy** | Baulac, Michel (2015); Furyk, Jeremy (2018) |
| **Feeding and Eating Disorders** | Davison, Karen M. (2017); Furth, Eric F. van (2016); Hart, Laura M. (2019) |
| **Intellectual Disability** | Johnson, Kelley (2014); Kramer, Jessica M. (2019); Tomlinson, Mark (2014); Tuffrey-Wijne, I. (2016) |
| **Learning Disabilities** | Lim, Ai Keow (2019); Paul, C. (2017) |
| **Neurodegenerative Diseases** | Iliffe, Steve (2013) |
| **Nutrition Therapy** | Davison, Karen M. (2017) |
| **Obsessive-Compulsive Disorder** | Kühne, F. (2019); McKay, Dean (2019) |
| **Other** | Brophy, Lisa (2018); Hitch, Danielle (2015) |
| **Patient Safety** | Dewa, Lindsay H. (2018) |
| **Psychological Distress** | Bell, Sigall K. (2018) |
| **Psychophysiologic Disorders** | van der Feltz-Cornelis, C.M. (2018); Zeigler, Vicki L. (2010) |
| **Schizophrenia** | Faulkner, Sophie (2017); McGurk, Susan R. (2013) |
| **Stroke** | Turner, Grace M. (2018) |
| **Substance-Related Disorders** | Clark, Kristen D. (2019); Kelber, Marija Spanovic (2019); Makeen, Anwar M. (2020) |
| **Suicide** | Booth, Chelsea L. (2014); Reifels, Lennart (2018); Roy, Kallol (2019) |
| **Tobacco Use Disorder** | Lindson, Nicola (2017); Makeen, Anwar M. (2020) |

Dementia is the most frequently studied specific condition, included in eleven articles followed by autistic disorder (7). Using the MeSH hierarchical organisation we can identify several major areas: neurodevelopmental disorders (autism, disabilities, attention disorder) are addressed by roughly one third of the studies (N=12), dementia (N=11), and neurobehavioral manifestations (intellectual and learning disabilities) (N=6).

Table S1, in the online Supplementary material, shows the disorders addressed by countries.

### Interest groups

We identified eleven interest groups participating in priority setting. The leading groups are practitioners (identified through textual expressions like “Paediatric neurologists”, “Emergency Physicians”, “Health professionals”, “Health and social care providers” and related terms) who participated in thirty-three original studies; public actors (community members, community/ public, parents, teachers, school counsellors, etc.) appear in twenty-six studies, and patients in twenty-four articles. Other groups include academics, experts (including legal professionals), policy makers, funders and service providers.

We classified the identified interest groups into five main categories: public (N=35), clinicians (N=34), researchers and academics (N=28), regulators (funders, representatives from organisms) (N=19), and other stakeholders (see Table S2 online Supplementary Material).

Table 3 shows clinicians interacted with all the other groups. Yet, more than one third of the studies (N=21) only included participants from a single group.

**Table 3.** Interest groups interactions

|  | Other | Regulators | Public | Clinicians | Researchers /Ac. |
| --- | --- | --- | --- | --- | --- |
| **R e s e a r c h e r s /**<br>**Academics** | 6 | 14 | 14 | 15 | 5 |
| **Clinicians** | 4 | 11 | 26 | 4 |  |
| **Public** | 3 | 11 | 9 |  |  |
| **Regulators** | 4 | 2 |  |  |  |
| **Other** | 1 |  |  |  |  |

### Methods, protocols and frameworks

Some studies used several techniques for requesting participants’ views while others use consensual criteria or accepted protocols to determine research priorities.

The dominant approach was the use of interviews and surveys (face-to-face and online). Workshops, meetings, and discussions were also common. Methods to reach decisions included consensus decision making processes (different Delphi variants were the most common, also James Lind Alliance protocols) and the nominal group technique. Online Table S4 contains a summary of the methods used. Literature reviews have been extensively used in combination with participative methods and authoritative resources. Table 4 shows the frequency of these associations. It is important to note the growing relevance of priority-setting partnerships (PSP), and the use of the James Lind Alliance protocols.

**Table 4.** Methods combined in the studies on priorities setting

|  | Theory of<br>change<br>methodology | Other | Priority<br>Setting<br>Partnership | Mixed<br>methods<br>research | Meetings<br>and<br>workshops | Literature<br>review | Interview | Group<br>decision-<br>making | Focus<br>group | Consensual<br>protocol |
| --- | --- | --- | --- | --- | --- | --- | --- | --- | --- | --- |
| Consensual<br>protocol |  |  | 1 |  | 1 | 1 | 2 |  |  |  |
| Focus<br>group | 1 | 2 |  | 1 |  | 1 | 9 | 1 |  |  |
| Group<br>decision-<br>making | 1 | 1 | 2 |  | 9 | 2 | 6 |  |  |  |
| Interview<br>and<br>consulting | 2 | 4 | 5 | 1 | 10 | 9 |  |  |  |  |
| Literature<br>review |  |  | 5 | 1 | 6 |  |  |  |  |  |
| Meetings<br>and<br>workshops |  |  | 5 |  |  |  |  |  |  |  |
| Mixed<br>methods<br>research |  |  | 1 |  |  |  |  |  |  |  |
| Priority<br>Setting<br>Partnership |  |  |  |  |  |  |  |  |  |  |
| Other | 1 |  |  |  |  |  |  |  |  |  |
| Theory of<br>change<br>methodology |  |  |  |  |  |  |  |  |  |  |

### Priorities identified

We transcribed, normalised and categorised 722 mental health research priorities using MeSH subheadings. Table 1 illustrates the classification process, Table S3 lists the subheadings used and their definitions, and Table S5 presents the links between categories’ subjects and their corresponding studies. On average, an academic article contains thirteen research priorities. The most comprehensive study[27] offers eighty-seven. The full list of the priorities is available upon request.

Overall (see Table 5 and Table 6) all interest groups emphasize the need for research on therapy, standards, education and psychology of mental disorders. The latter includes the psycho-social aspects of the diseases. Nonetheless, different interest group categories favour different priorities: while therapy, diagnosis, methods and standards-related priorities rank highly among clinicians, public and researchers, the latter differentiate from the others in significant priorities. Scientists generally do not rank high education-related priorities, whereas these are well positioned among the priorities preferred by clinicians and public, and the same applies to rehabilitation- and complications-related priorities.

**Table 5.** Priorities selected by the stakeholders.

| Research priorities group | Clinicians | Public | Regulators | Researchers/Ac. | Other |
| --- | --- | --- | --- | --- | --- |
| classification | 1,93% | 1,73% | 1,82% | 2,58% | 0% |
| complications | 5,80% | 5,19% | 1,82% | 5,16% | 6,52% |
| diagnosis | 5,31% | 6,49% | 4,55% | 7,10% | 4,35% |
| diagnostic imaging | 0,48% | 0,43% | 0,91% | 0% | 0% |
| diet therapy | 0,97% | 0,87% | 0,91% | 0,65% | 2,17% |
| drug therapy | 2,42% | 4,76% | 0,91% | 1,94% | 0% |
| economics | 3,38% | 3,46% | 3,64% | 3,23% | 0% |
| education | 9,66% | 9,96<br>% | 6,36% | 5,81% | 6,52% |
| epidemiology | 2,42% | 1,30<br>% | 5,45% | 2,58% | 2,17% |
| ethnology | 1,45% | 1,73<br>% | 4,55% | 2,58% | 4,35% |
| etiology | 3,86% | 3,46<br>% | 3,64% | 4,52% | 6,52% |
| genetics | 1,45% | 1,30<br>% | 0,91% | 0,65% | 0% |
| l e g i s l a t i o n &<br>jurisprudence | 0,97% | 1,30<br>% | 0,91% | 1,29% | 0% |
| methods | 6,76% | 6,49<br>% | 10,00% | 9,03% | 6,52% |
| o r g a n i z a t i o n &<br>administration | 2,90% | 3,46<br>% | 4,55% | 4,52% | 0% |
| physiopathology | 2,90% | 2,16<br>% | 3,64% | 2,58% | 0% |
| prevention & control | 5,80% | 6,06<br>% | 9,09% | 7,10% | 13,04% |
| psychology | 7,25% | 7,36<br>% | 9,09% | 7,74% | 15,22% |
| rehabilitation | 7,25% | 6,93<br>% | 2,73% | 3,87% | 6,52% |
| standards | 8,70% | 7,79<br>% | 9,09% | 9,03% | 10,87% |
| s t a t i s t i c s &<br>numerical data | 0,97% | 0,87<br>% | 0,91% | 0,65% | 0% |
| supply & distribution | 7,25% | 6,93<br>% | 6,36% | 8,39% | 8,70% |
| therapy | 10,14% | 9,96<br>% | 8,18% | 9,03% | 6,52% |

**Table 6.**
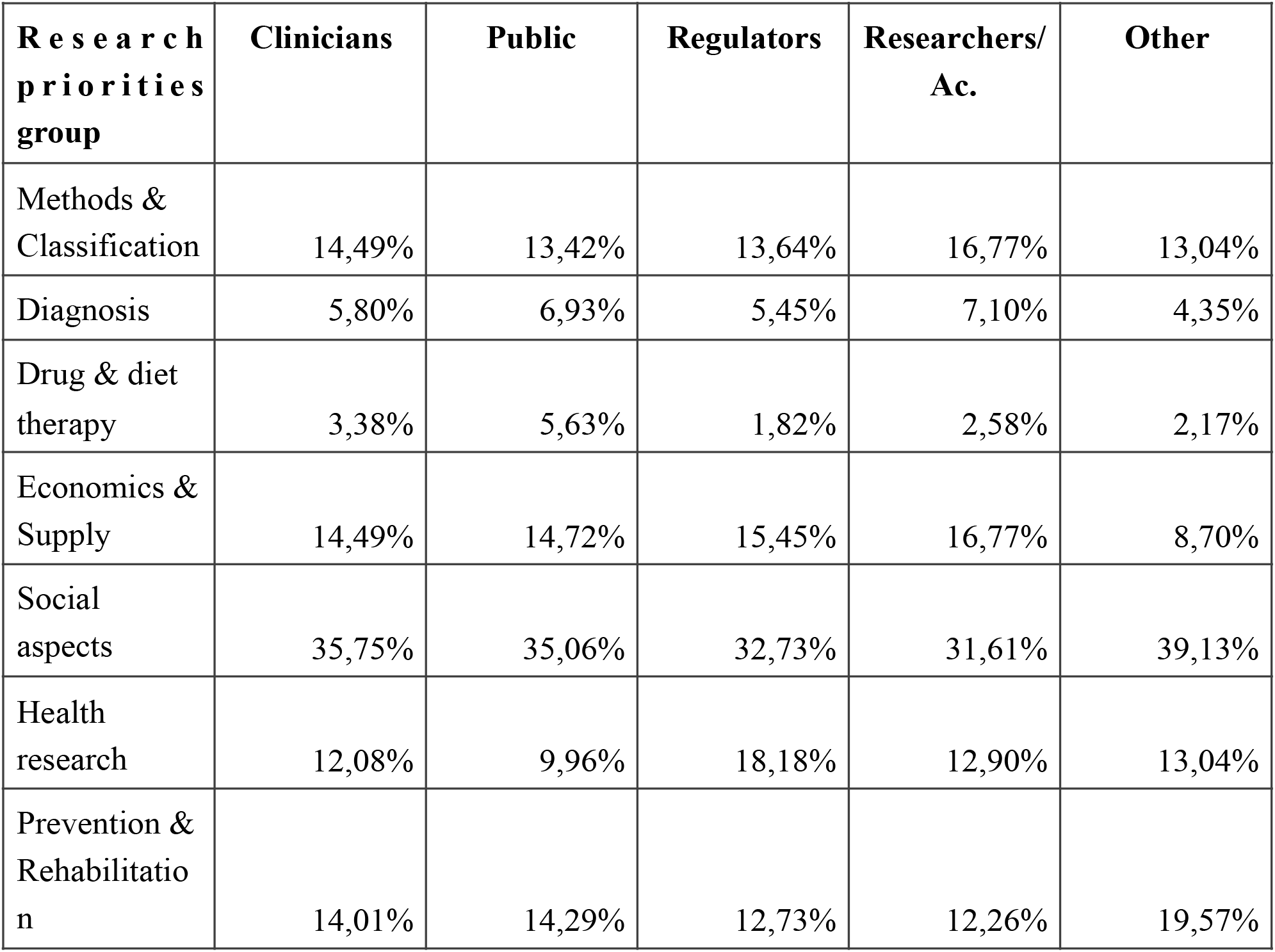
Priority clusters selected by the stakeholders.

In cluster analysis (Table 6) we merged in seven groups those priorities related to each other. In group one (Methods & classification) we include classification, complications and methods; in group two (Diagnosis) we include diagnosis and diagnostic imaging; in group three (Drug and diet therapy) it is included diet therapy and drug therapy; in group four (Economics & supply) we include economics, organization & administration, statistics & numerical data, and supply & distribution; in group five (Social aspects) it is included education, psychology, standards, and therapy; in group six (Health research) we included epidemiology, ethnology, etiology, genetics, and physiopathology; finally in group seven (Prevention & Rehabilitation) we included legislation & jurisprudence, prevention & control and rehabilitation. As shown, priorities are well-distributed among all interest groups. It is interesting to note that researchers do not prioritise social aspects as clinicians and public; scientists prioritise more the methodological aspects of mental disorders.

## Conclusion

The scoping review results on the participation of social groups in priority setting in mental disorders research show that different groups prioritise different health areas.

Scientists differentiate themselves from practitioners, the public (patients, communities) and organisations in not giving preference to education-related and rehabilitation research priorities. In addition, groups do not extensively collaborate with each other, more than one third of the studies only included participants from a single interest group. Patients and communities (i.e. “public” category) participated in most of the priority setting studies reviewed. Generally, those among the public category collaborate with clinicians in designing the studies, although there is no evidence suggesting public involvement in the outcomes evaluation. We noted that approaches combining a participatory strategy with literature reviews, consensual protocols and authoritative sources are common.

We can conclude priority setting partnerships are becoming more popular, with the use of the James Lind Alliance protocols gaining momentum. This can be potentially extended to other participatory activities in priority setting across different countries. However, according to our results, whereas participatory methods are spreading, the geographical scope of the studies is not well distributed and it lacks international collaboration: English speaking countries and the Netherlands concentrate the majority of the original studies, two thirds of which are produced by authors from a single country. In health areas, we have seen neurodevelopmental disorders and dementia, followed by neurobehavioral manifestations, are the most common mental disorders tackled by participatory priority setting methods.

Scoping reviews are recommended for examining the extent, variety and nature of an ill-defined and broad research topic, and they are considered a previous step to eventually performing more resourceful synthesis studies. In view of our results, it seems a logical follow up to develop a systematic review through literature on priority setting in mental disorders research

## Supporting information

List of sources of evidence

## Data Availability

All data produced in the present work are provided in the supplementary material accompanying the manuscrpit

## Acknowledgments

This study was partially supported by the Spanish Ministry of Science and Innovation under project MAPAMENT (PRE2018-084855).

## Footnotes

3 Users, as commonly seen in literature, refers to citizens who benefit from research results, i.e. not only patients.

