## Supplementary material for "Priorities setting in mental health research: a scoping review": List of sources of evidence

### List of sources of evidence for MAPAMENT Study

Entries marked with an asterisk are non-original studies.

1.
Banfield MA et al. Australian mental health consumers' priorities for research: Qualitative findings from the SCOPE for Research project. Health Expectations [Internet]. 2014 [citado 3 de febrero de 2020];17(3):365-75. Disponible en: https://onlinelibrary.wiley.com/doi/abs/10.1111/j.1369-7625.2011.00763.x
2.
Banfield MA et al. SCOPE for Research: mental health consumers' priorities for research compared with recent research in Australia. Australian and New Zealand Journal of Psychiatry [Internet]. 1 de diciembre de 2011 [citado 3 de febrero de 2020];45(12):1078-85. Disponible en: https://www.tandfonline.com/doi/abs/10.3109/00048674.2011.624084
3.
Banfield MA et al. Mental health research priorities in Australia: a consumer and carer agenda. Health Res Policy Syst [Internet]. 12 de diciembre de 2018 [citado 3 de febrero de 2020];16. Disponible en: https://www.ncbi.nlm.nih.gov/pmc/articles/PMC6292010/
4.
* Barry MM, Clarke AM, Petersen I. Promotion of mental health and prevention of mental disorders: priorities for implementation. East Mediterr Health J. 28 de septiembre de 2015;21(7):503-11.
5.
Baulac M et al. Epilepsy priorities in Europe: A report of the ILAE−IBE Epilepsy Advocacy Europe Task Force. Epilepsia [Internet]. noviembre de 2015 [citado 3 de febrero de 2020];56(11):1687-95. Disponible en: https://www.ncbi.nlm.nih.gov/pmc/articles/PMC5019256/
6.
Bell SK et al. A Multi-Stakeholder Consensus-Driven Research Agenda for Better Understanding and Supporting the Emotional Impact of Harmful Events on Patients and Families. The Joint Commission Journal on Quality and Patient Safety [Internet]. 1 de julio de 2018 [citado 3 de febrero de 2020];44(7):424-35. Disponible en: http://www.sciencedirect.com/science/article/pii/S1553725018300540
7.
Bethell J et al. Patient and Public Involvement in Identifying Dementia Research Priorities. Journal of the American Geriatrics Society [Internet]. 2018 [citado 3 de febrero de 2020];66(8):1608-12. Disponible en: https://onlinelibrary.wiley.com/doi/abs/10.1111/jgs.15453
8.

* Bigby C, Frawley P, Ramcharan P. A collaborative group method of inclusive research. Journal of Applied Research in Intellectual Disabilities [Internet]. enero de 2014;27(1):54-64. Disponible en: https://search.proquest.com/docview/1499090707?accountid=14777
9.
Blum NJ et al. Research priorities for developmental-behavioral pediatrics: a DBPNet consensus study. J Dev Behav Pediatr. julio de 2012;33(6):509-16.
10.
* Booth CL. Experiences and Wisdom Behind the Numbers: Qualitative Analysis of the National Action Alliance for Suicide Prevention's Research Prioritization Task Force Stakeholder Survey. American Journal of Preventive Medicine [Internet]. 1 de septiembre de 2014 [citado 27 de mayo de 2020];47(3):S106-14. Disponible en: https://www.ajpmonline.org/article/S0749-3797(14)00240-2/abstract
11.
Brophy L et al. Community treatment orders: towards a new research agenda. Australas Psychiatry [Internet]. 1 de junio de 2018 [citado 3 de febrero de 2020]; 26(3):299-302. Disponible en: https://doi.org/10.1177/1039856218758543
12.
Camden C et al. Research and knowledge transfer priorities in developmental coordination disorder: Results from consultations with multiple stakeholders. Health Expectations: An International Journal of Public Participation in Health Care & Health Policy [Internet]. 14 de agosto de 2019; Disponible en: https://search.proquest.com/docview/2273353221?accountid=14777
13.
Clark KD et al. Supporting sexual and gender minority health: Research priorities from mental health professionals. Journal of Gay and Lesbian Mental Health [Internet]. 2020;24(2):205-21. Disponible en: https://www.scopus.com/inward/record.uri?eid=2-s2.0-85076879681&doi=10.1080%2f19359705.2019.1700865&partnerID=40&md5=25692b3455a2b9bd3423c7cd9e607392
14.
Clark M, Adams D. Listening to parents to understand their priorities for autism research. PLoS One. 2020;15(8):e0237376.
15.
Clavisi O et al. Effective stakeholder participation in setting research priorities using a Global Evidence Mapping approach. J Clin Epidemiol. mayo de 2013;66(5): 496-502.e2.
16.

* Collins M et al. Using the Public Involvement Impact Assessment Framework to assess the impact of public involvement in a mental health research context: A reflective case study. Health Expect. 2018;21(6):950-63.
17.
Crocker AG et al. Research Priorities in Mental Health, Justice, and Safety: A Multidisciplinary Stakeholder Report. International Journal of Forensic Mental Health [Internet]. 3 de julio de 2015 [citado 3 de febrero de 2020];14(3):205-17. Disponible en: https://doi.org/10.1080/14999013.2015.1073197
18.
Davison KM et al. The development of a national nutrition and mental health research agenda with comparison of priorities among diverse stakeholders. Public Health Nutrition [Internet]. marzo de 2017 [citado 3 de febrero de 2020];20(4):712-25. Disponible en: https://www.cambridge.org/core/journals/public-health-nutrition/article/development-of-a-national-nutrition-and-mental-health-research-agenda-with-comparison-of-priorities-among-diverse-stakeholders/9D2E17D4F15F1315D13A53EC007EC5BD
19.
* Daya I, Hamilton B, Roper C. Authentic engagement: A conceptual model for welcoming diverse and challenging consumer and survivor views in mental health research, policy, and practice. Int J Ment Health Nurs. 20 de septiembre de 2019;
20.
Elfeddali I et al. Horizon 2020 Priorities in Clinical Mental Health Research: Results of a Consensus-Based ROAMER Expert Survey. Int J Environ Res Public Health [Internet]. octubre de 2014 [citado 3 de febrero de 2020];11(10):10915-39. Disponible en: https://www.ncbi.nlm.nih.gov/pmc/articles/PMC4211014/
21.
* Faithfull S et al. Barriers and enablers to meaningful youth participation in mental health research: qualitative interviews with youth mental health researchers. J Ment Health. febrero de 2019;28(1):56-63.
22.
Fiorillo A et al. Priorities for mental health research in Europe: A survey among national stakeholders' associations within the ROAMER project. World Psychiatry. junio de 2013;12(2):165-70.
23.
Forsman AK et al. Research priorities for public mental health in Europe: recommendations of the ROAMER project. Eur J Public Health [Internet]. 1 de abril de 2015 [citado 3 de febrero de 2020];25(2):249-54. Disponible en: https://academic.oup.com/eurpub/article/25/2/249/491448
24.

Furth EF van, Meer A van der, Cowan K. Top 10 research priorities for eating disorders. The Lancet Psychiatry [Internet]. 1 de agosto de 2016 [citado 3 de febrero de 2020];3(8):706-7. Disponible en: https://www.thelancet.com/journals/lanpsy/article/PIIS2215-0366(16)30147-X/abstract
25.
Furyk J et al. Consensus research priorities for paediatric status epilepticus: A Delphi study of health consumers, researchers and clinicians. Seizure [Internet]. marzo de 2018;56:104-9. Disponible en: https://search.proquest.com/docview/2130160909?accountid=14777
26.
Gaynes BN et al. Attention-Deficit/Hyperactivity Disorder: Identifying high priority future research needs. Journal of Psychiatric Practice [Internet]. marzo de 2014;20(2):104-17. Disponible en: https://search.proquest.com/docview/1548788302?accountid=14777
27.
Ghisoni M et al. Priority setting in research: user led mental health research. Res Involv Engagem. 2017;3:4.
28.
* Giovino GA et al. Research Priorities for FCTC Articles 20, 21, and 22: Surveillance/Evaluation and Information Exchange. Nicotine Tob Res. abril de 2013;15(4):847-61.
29.
Glandon D et al. Identifying health policy and systems research priorities on multisectoral collaboration for health in low-income and middle-income countries. BMJ Glob Health. 2018;3(Suppl 4):e000970.
30.
* Goold SD et al. Evaluating community deliberations about health research priorities. Health Expectations: An International Journal of Public Participation in Health Care & Health Policy [Internet]. 28 de junio de 2019; Disponible en: https://search.proquest.com/docview/2249978559?accountid=14777
31.
Gregório G et al. Setting priorities for mental health research in Brazil. Brazilian Journal of Psychiatry [Internet]. diciembre de 2012 [citado 3 de febrero de 2020];34(4):434-9. Disponible en: http://www.scielo.br/scielo.php?script=sci_abstract&pid=S1516-44462012000400010&lng=en&nrm=iso&tlng=en
32.
Hart LM, Wade T. Identifying research priorities in eating disorders: A delphi study building consensus across clinicians, researchers, consumers, and carers in australia. International Journal of Eating Disorders [Internet]. 30 de septiembre de 2019; Disponible en: https://search.proquest.com/docview/2300267488?accountid=14777

33.
Hitch D, Lhuede K. Research priorities in mental health occupational therapy: A study of clinician perspectives. Australian Occupational Therapy Journal. 2015;62(5): 326-32.
34.
Hollis C et al. Identifying research priorities for digital technology in mental health care: results of the James Lind Alliance Priority Setting Partnership. The Lancet Psychiatry [Internet]. 2018;5(10):845-54. Disponible en: https://www.scopus.com/inward/record.uri?eid=2-s2.0-85054049876&doi=10.1016%2fS2215-0366%2818%2930296-7&partnerID=40&md5=968895ada0d91d47625d637c44f71771
35.
Jacobson S et al. TOP TEN RESEARCH PRIORITIES FOR ATTENTION DEFICIT/HYPERACTIVITY DISORDER TREATMENT. International Journal of Technology Assessment in Health Care [Internet]. ed de 2016 [citado 3 de febrero de 2020];32(3): 152-9. Disponible en: https://www.cambridge.org/core/journals/international-journal-of-technology-assessment-in-health-care/article/top-ten-research-priorities-for-attention-deficithyperactivity-disorder-treatment/AA8E6B6E5C7DCAB7132CC526FC397504
36.
* Johnson B. Schizophrenia research priorities from the viewpoint of patients/relatives. Die Psychiatrie: Grundlagen & Perspektiven [Internet]. 2016;13(3):161-2. Disponible en: https://search.proquest.com/docview/1901541610?accountid=14777
37.
Kelber MS et al. Identifying research gaps in substance use disorder: A systematic methodology and prioritized list. Am J Drug Alcohol Abuse. 4 de julio de 2019;45(4): 355-64.
38.
Kelly S et al. Dementia priority setting partnership with the James Lind Alliance: using patient and public involvement and the evidence base to inform the research agenda. Age Ageing [Internet]. noviembre de 2015 [citado 3 de febrero de 2020]; 44(6):985-93. Disponible en: https://www.ncbi.nlm.nih.gov/pmc/articles/PMC4621237/
39.
Khandelwal S et al. Mental and neurological health research priorities setting in developing countries. Soc Psychiat Epidemiol [Internet]. 1 de abril de 2010 [citado 7 de febrero de 2020];45(4):487-95. Disponible en: https://doi.org/10.1007/s00127-009-0089-2
40.

Kramer JM et al. Improving research and practice: Priorities for young adults with intellectual/developmental disabilities and mental health needs. Journal of Mental Health Research in Intellectual Disabilities [Internet]. 2 de agosto de 2019; Disponible en: https://search.proquest.com/docview/2269454955?accountid=14777
41.
Kühne F et al. Research priorities set by people with OCD and OCD researchers: Do the commonalities outweigh the differences? Health Expectations. 2019;
42.
Law E, Starr JM, Connelly PJ. Dementia research – what do different public groups want? A survey by the Scottish Dementia Clinical Research Network. Dementia [Internet]. 1 de enero de 2013 [citado 3 de febrero de 2020];12(1):23-8. Disponible en: https://doi.org/10.1177/1471301211420309
43.
Lee C et al. Identifying research priorities for psychosocial support programs in humanitarian settings. Glob Ment Health (Camb). 2019;6:e23.
44.
Leroi I et al. A roadmap to develop dementia research capacity and capability in Pakistan: A model for low- and middle-income countries. Alzheimers Dement (N Y). 2019;5:939-52.
45.
Lim AK et al. Joint production of research priorities to improve the lives of those with childhood onset conditions that impair learning: the James Lind Alliance Priority Setting Partnership for «learning difficulties». BMJ Open. 30 de octubre de 2019;9(10):e028780.
46.
Lindson N et al. Setting research priorities in tobacco control: a stakeholder engagement project. Addiction. diciembre de 2017;112(12):2257-71.
47.
Maassen EF et al. A research agenda for bipolar disorder developed from a patients' perspective. Journal of Affective Disorders [Internet]. 15 de octubre de 2018;239:11-7. Disponible en: https://search.proquest.com/docview/2118090737?accountid=14777
48.
* MacGabhann L et al. Democratic communities: evaluating trialogue for mental health stakeholders. Ment Health Rev J. 2018;23(2):94-109.
49.
Makeen AM et al. Delphi consensus on research priorities in tobacco use and substance abuse in Saudi Arabia. J Ethn Subst Abuse. 2020;
50.

* McRobbie H, Raw M, Chan S. Research Priorities for Article 14-Demand Reduction Measures Concerning Tobacco Dependence and Cessation. Nicotine Tob Res. abril de 2013;15(4):805-16.
51.
Mei C et al. Global research priorities for youth mental health. Early Intervention in Psychiatry [Internet]. 2020 [citado 3 de febrero de 2020];14(1):3-13. Disponible en: https://onlinelibrary.wiley.com/doi/abs/10.1111/eip.12878
52.
* Miah J et al. Patient and public involvement in dementia research in the European Union: a scoping review. BMC Geriatr. 14 de agosto de 2019;19(1):220.
53.
* Murphy JK et al. Methodological approaches to situational analysis in global mental health: a scoping review. Global Mental Health [Internet]. ed de 2019 [citado 20 de marzo de 2020];6. Disponible en: https://www.cambridge.org/core/journals/global-mental-health/article/methodological-approaches-to-situational-analysis-in-global-mental-health-a-scoping-review/CB5F23FCBF3AA721C4921B86E090DCDE
54.
* Nagler RH, Viswanath K. Implementation and Research Priorities for FCTC Articles 13 and 16: Tobacco Advertising, Promotion, and Sponsorship and Sales to and by Minors. Nicotine Tob Res. abril de 2013;15(4):832-46.
55.
Obeid N et al. Cocreating research priorities for anorexia nervosa: The canadian eating disorder priority setting partnership. International Journal of Eating Disorders [Internet]. 3 de febrero de 2020; Disponible en: https://search.proquest.com/docview/2351606534?accountid=14777
56.
* Paul C, Holt J. Involving the public in mental health and learning disability research: Can we, should we, do we? J Psychiatr Ment Health Nurs. octubre de 2017;24(8):570-9.
57.
Pellicano E, Dinsmore A, Charman T. What should autism research focus upon? Community views and priorities from the United Kingdom. Autism [Internet]. octubre de 2014;18(7):756-70. Disponible en: https://search.proquest.com/docview/1707075150?accountid=14777
58.
Pickett J et al. A roadmap to advance dementia research in prevention, diagnosis, intervention, and care by 2025. Int J Geriatr Psychiatry [Internet]. julio de 2018 [citado 3 de febrero de 2020];33(7):900-6. Disponible en: https://www.ncbi.nlm.nih.gov/pmc/articles/PMC6033035/
59.

Reifels L et al. Research Priorities in Suicide Prevention: Review of Australian Research from 2010–2017 Highlights Continued Need for Intervention Research. Int J Environ Res Public Health. 20 de 2018;15(4).
60.
* Russell G et al. Selective patient and public involvement: The promise and perils of pharmaceutical intervention for autism. Health Expect [Internet]. abril de 2018 [citado 3 de febrero de 2020];21(2):466-73. Disponible en: https://www.ncbi.nlm.nih.gov/pmc/articles/PMC5867326/
61.
Shah H et al. Research priorities to reduce the global burden of dementia by 2025. The Lancet Neurology [Internet]. 1 de noviembre de 2016 [citado 3 de febrero de 2020];15(12):1285-94. Disponible en: http://www.sciencedirect.com/science/article/pii/S1474442216302356
62.
Shattuck PT et al. A National Research Agenda for the Transition of Youth With Autism. Pediatrics [Internet]. 1 de abril de 2018 [citado 3 de febrero de 2020]; 141(Supplement 4):S355-61. Disponible en: https://pediatrics.aappublications.org/content/141/Supplement_4/S355
63.
* Stein DJ. Psychiatry and mental health research in South Africa: National priorities in a low and middle income context. African Journal of Psychiatry [Internet]. 1 de enero de 2012 [citado 3 de febrero de 2020];15(6):427-431-431. Disponible en: https://www.ajol.info/index.php/ajpsy/article/view/83483
64.
* Tapsell A et al. Expert by Experience Involvement in Mental Health Research: Developing a Wellbeing Brochure for People with Lived Experiences of Mental Illness. Issues Ment Health Nurs. 9 de enero de 2020;1-7.
65.
Tol WA et al. Relevance or Excellence? Setting Research Priorities for Mental Health and Psychosocial Support in Humanitarian Settings. Harvard Review of Psychiatry [Internet]. 8 de febrero de 2012 [citado 3 de febrero de 2020];20(1):25-36. Disponible en: https://www.tandfonline.com/doi/abs/10.3109/10673229.2012.649113
66.
Tomlinson M et al. Setting global research priorities for developmental disabilities, including intellectual disabilities and autism. Journal of Intellectual Disability Research [Internet]. 2014 [citado 3 de febrero de 2020];58(12):1121-30. Disponible en: https://onlinelibrary.wiley.com/doi/abs/10.1111/jir.12106
67.
Tuffrey-Wijne I et al. Developing research priorities for palliative care of people with intellectual disabilities in Europe: a consultation process using nominal group

technique. BMC Palliat Care [Internet]. 24 de marzo de 2016 [citado 3 de febrero de 2020];15. Disponible en: https://www.ncbi.nlm.nih.gov/pmc/articles/PMC4806426/
68.
Turner GM et al. Establishing research priorities relating to the long-term impact of TIA and minor stroke through stakeholder-centred consensus. Res Involv Engagem [Internet]. 25 de enero de 2018 [citado 3 de febrero de 2020];4. Disponible en: https://www.ncbi.nlm.nih.gov/pmc/articles/PMC5784709/
69.
van der Feltz-Cornelis CM et al. A European research agenda for somatic symptom disorders, bodily distress disorders, and functional disorders: Results of an estimate-talk-estimate delphi expert study. Frontiers in Psychiatry [Internet]. 2018;9(MAY). Disponible en: https://www.scopus.com/inward/record.uri?eid=2-s2.0-85047003409&doi=10.3389%2ffpsyt.2018.00151&partnerID=40&md5=2ff3fca4cd9bdeb5d5382f3f5a7fcd88
70.
van der Feltz-Cornelis CM et al. Towards Horizon 2020: challenges and advances for clinical mental health research – outcome of an expert survey. Neuropsychiatr Dis Treat [Internet]. 27 de junio de 2014 [citado 3 de febrero de 2020];10:1057-68. Disponible en: https://www.ncbi.nlm.nih.gov/pmc/articles/PMC4085314/
71.
Vasa RA et al. Priorities for advancing research on youth with autism spectrum disorder and co-occurring anxiety. Journal of Autism and Developmental Disorders [Internet]. marzo de 2018;48(3):925-34. Disponible en: https://search.proquest.com/docview/1968546070?accountid=14777
72.
* Weiss WM et al. Mental health interventions and priorities for research for adult survivors of torture and systematic violence: a review of the literature. Torture. 2016;26(1):17-44.
73.
Wykes T et al. Mental health research priorities for Europe. The Lancet Psychiatry [Internet]. 1 de noviembre de 2015 [citado 3 de febrero de 2020];2(11):1036-42. Disponible en: http://www.sciencedirect.com/science/article/pii/S2215036615003326
74.
Zitko P et al. Priority setting for mental health research in Chile. Int J Ment Health Syst. 2017;11:61.
