## Supplementary tables for "Priorities setting in mental health research: a scoping review"

#### Table RRR. Mental disorders covered by the articles

| | |
|---|---|
| Alzheimer Disease | Gove, Dianne (2018); Liggins, Charlene (2014) |
| Attention Deficit Disorder with Hyperactivity | Obeid, Nicole (2020) |
| Autistic Disorder | Frazier, Thomas W. (2018); Pellicano, Elizabeth (2014); Russell, Ginny (2018); Shattuck, Paul T. (2018); Tomlinson, Mark (2014); Vasa, Roma A. (2018) |
| Bipolar Disorder | Banfield, Michelle A. (2011); Banfield, Michelle A. (2014); Maassen, Eva F. (2018) |
| Brain Injuries, Traumatic | Clavisi, Ornella (2013) |
| Delirium | Pandharipande, Pratik (2017) |
| Dementia | Bethell, Jennifer (2018); Iliffe, Steve (2013); Kelly, Sarah (2015); Law, Emma (2013); Leroi, Iracema (2019); Miah, Jahanara (2019); Pickett, James (2018); Schneider, Lon S. (2016); Shah, Hiral (2016) |
| Depressive Disorder | Banfield, Michelle A. (2011); Banfield, Michelle A. (2014); Topooco, Naira (2017) |
| Developmental Disabilities | Blum, Nathan J. (2012); Camden, Chantal (2019); Kramer, Jessica M. (2019) |
| Dyssomnias | Bassetti, C. L. (2015) |
| Epilepsy | Baulac, Michel (2015); Furyk, Jeremy (2018) |
| Feeding and Eating Disorders | Davison, Karen M. (2017); Furth, Eric F. van (2016); Hart, Laura M. (2019) |
| Intellectual Disability | Johnson, Kelley (2014); Kramer, Jessica M. (2019); Tomlinson, Mark (2014); Tuffrey-Wijne, I. (2016) |
| Learning Disabilities | Lim, Ai Keow (2019); Paul, C. (2017) |
| Neurodegenerative Diseases | Iliffe, Steve (2013) |
| Obsessive-Compulsive Disorder | Kühne, F. (2019); McKay, Dean (2019) |
| Psychological Distress | Bell, Sigall K. (2018) |
| Psychophysiologic Disorders | van der Feltz-Cornelis, C.M. (2018); Zeigler, Vicki L. (2010) |
| Schizophrenia | McGurk, Susan R. (2013) |
| Stroke | Turner, Grace M. (2018) |
| Substance-Related Disorders | Clark, Kristen D. (2019) |
| Suicide | Booth, Chelsea L. (2014); Reifels, Lennart (2018); Roy, Kallol (2019) |

### Table ZZ. Country contribution to research priorities setting in mental disorders

| country | # papers | references |
|---|---|---|
| Australia | 18 | Clavisi, Ornella (2013); Tapsell, Amy (2020); Furyk, Jeremy (2018); Mei, Cristina (2020); Banfield, Michelle A. (2011); Tomlinson, Mark (2014); Faithfull, Sarah (2019); Murphy, J. K. (2019); Hitch, Danielle (2015); Brophy, Lisa (2018); Khandelwal, Sudhir (2010); Finan, Samantha Jane (2020); Tol, Wietse A. (2012); Shah, Hiral (2016); Reifels, Lennart (2018); Hart, Laura M. (2019); Banfield, Michelle A. (2018); Banfield, Michelle A. (2014) |
| Austria | 2 | Forsman, Anna K. (2015); MacGabhann, Liam (2018) |
| Belgium | 1 | van der Feltz-Cornelis, C.M. (2018) |
| Benin | 1 | Khandelwal, Sudhir (2010) |
| Brazil | 1 | Gregório, Guilherme (2012) |
| Brunei | 1 | Khandelwal, Sudhir (2010) |
| Bulgaria | 1 | Khandelwal, Sudhir (2010) |
| Canada | 9 | Davison, Karen M. (2017); Mei, Cristina (2020); Tomlinson, Mark (2014); Crocker, Anne G. (2015); Bethell, Jennifer (2018); Murphy, J. K. (2019); Obeid, Nicole (2020); Shah, Hiral (2016); Camden, Chantal (2019) |
| Chile | 2 | Khandelwal, Sudhir (2010); Zitko, Pedro (2017) |
| China | 1 | Shah, Hiral (2016) |
| Denmark | 1 | Russell, Ginny (2018) |
| Deutschland | 10 | Kühne, F. (2019); Forsman, Anna K. (2015); Tuffrey-Wijne, I. (2016); van der Feltz-Cornelis, C.M. (2018); Elfeddali, Iman (2014); Wykes, Til (2015); Baulac, Michel (2015); Hazo, Jean-Baptiste (2019); Khandelwal, Sudhir (2010); van der Feltz-Cornelis, Christina M (2014) |
| Ecuador | 1 | Khandelwal, Sudhir (2010) |
| Egipt | 1 | Khandelwal, Sudhir (2010) |
| England | 28 | Dewa, Lindsay H. (2018); Parikh, R. (2019); Forsman, Anna K. (2015); Jennings, Helen (2018); Tuffrey-Wijne, I. (2016); Mei, Cristina (2020); Tomlinson, Mark (2014); van der Feltz-Cornelis, C.M. (2018); Bethell, Jennifer (2018); Lim, Ai Keow (2019); Furth, Eric F. van (2016); Paul, C. (2017); Wykes, Til (2015); Collins, Michelle (2018); Kearney, Anna (2017); Kelly, Sarah (2015); Hazo, Jean-Baptiste (2019); Khandelwal, Sudhir (2010); Pellicano, Elizabeth (2014); Tol, Wietse A. (2012); Shah, Hiral (2016); Turner, Grace M. (2018); van der Feltz-Cornelis, Christina M (2014); Hollis, Chris (2018); Miah, Jahanara (2019); Leroi, Iracema (2019); Russell, Ginny (2018); Pickett, James (2018) |
| Estonia | 1 | Forsman, Anna K. (2015) |
| Philippines | 1 | Khandelwal, Sudhir (2010) |
| Finland | 5 | Forsman, Anna K. (2015); Wykes, Til (2015); Baulac, Michel (2015); Hazo, Jean-Baptiste (2019); van der Feltz-Cornelis, Christina M (2014) |

| | | |
|---|---|---|
| France | 5 | Forsman, Anna K. (2015); Wykes, Til (2015); Baulac, Michel (2015); Hazo, Jean-Baptiste (2019); Khandelwal, Sudhir (2010) |
| Hungary | 1 | Wykes, Til (2015) |
| Iceland | 1 | Forsman, Anna K. (2015) |
| India | 3 | Parikh, R. (2019); Khandelwal, Sudhir (2010); Shah, Hiral (2016) |
| Ireland | 6 | Forsman, Anna K. (2015); Tuffrey-Wijne, I. (2016); Mei, Cristina (2020); MacGabhann, Liam (2018); Baulac, Michel (2015); Leroi, Iracema (2019) |
| Italy | 4 | Forsman, Anna K. (2015); Wykes, Til (2015); Baulac, Michel (2015); Fiorillo, Andrea (2013) |
| Kenia | 1 | Khandelwal, Sudhir (2010) |
| Lithuania | 1 | Khandelwal, Sudhir (2010) |
| Malta | 1 | Baulac, Michel (2015) |
| Myanmar | 1 | Parikh, R. (2019) |
| Netherlands | 13 | Baulac, Michel (2015); Elfeddali, Iman (2014); Forsman, Anna K. (2015); Furth, Eric F. van (2016); Hazo, Jean-Baptiste (2019); Khandelwal, Sudhir (2010); Maassen, Eva F. (2018); Parikh, R. (2019); Tol, Wietse A. (2012); Tuffrey-Wijne, I. (2016); van der Feltz-Cornelis, C.M. (2018); van der Feltz-Cornelis, Christina M (2014); Wykes, Til (2015) |
| New Zealand | 1 | Furyk, Jeremy (2018) |
| North Ireland | 1 | Kearney, Anna (2017) |
| Norway | 2 | Forsman, Anna K. (2015); van der Feltz-Cornelis, C.M. (2018) |
| Pakistan | 1 | Leroi, Iracema (2019) |
| Portugal | 1 | Forsman, Anna K. (2015) |
| Scotland | 2 | Law, Emma (2013); Lim, Ai Keow (2019) |
| South Africa | 3 | Gregório, Guilherme (2012); Tol, Wietse A. (2012); Tomlinson, Mark (2014) |
| Spain | 7 | Elfeddali, Iman (2014); Fiorillo, Andrea (2013); Forsman, Anna K. (2015); Hazo, Jean-Baptiste (2019); van der Feltz-Cornelis, C.M. (2018); van der Feltz-Cornelis, Christina M (2014); Wykes, Til (2015) |
| Sri Lanka | 1 | Tol, Wietse A. (2012) |
| Sweden | 5 | Forsman, Anna K. (2015); Jacobson, Stella (2016); Tuffrey-Wijne, I. (2016); van der Feltz-Cornelis, C.M. (2018); Wykes, Til (2015) |
| Switzerland | 6 | Baulac, Michel (2015); Shah, Hiral (2016); Tol, Wietse A. (2012); Tomlinson, Mark (2014); Tuffrey-Wijne, I. (2016); van der Feltz-Cornelis, C.M. (2018) |
| Uganda | 2 | Khandelwal, Sudhir (2010); Tol, Wietse A. (2012) |
| USA | 17 | Bell, Sigall K. (2018); Bethell, Jennifer (2018); Blum, Nathan J. (2012); Clark, Kristen D. (2019); Gaynes, Bradley N. (2014); Glandon, Douglas (2018); Jonas, Daniel E. (2012); Khandelwal, Sudhir (2010); Kramer, Jessica M. (2019); Lee, C. (2019); Mei, Cristina (2020); Murphy, J. K. (2019); Parikh, R. (2019); Shah, Hiral (2016); Shattuck, Paul T. (2018); Tol, Wietse A. (2012); Vasa, Roma A. (2018) |

| | | |
|---|---|---|
| Wales | 3 | Ghisoni, Marjorie (2017); Kearney, Anna (2017); Pickett, James (2018) |
| Zambia | 1 | Khandelwal, Sudhir (2010) |

#### Table FFF. Techniques used in gathering the opinion of the stakeholders

| techniques | textual expressions | Papers |
|---|---|---|
| **Consensual protocol** | Child Health and Nutrition Research Initiative Priority Setting; PICO framework; European Forum on Epilepsy Research ; combined approach matrix of the GNRMNH | Tomlinson, Mark (2014); Kelly, Sarah (2015); Baulac, Michel (2015); Khandelwal, Sudhir (2010); Shah, Hiral (2016) |
| **Focus group** | focus groups; community forums | Pellicano, Elizabeth (2014); Tol, Wietse A. (2012); Leroi, Iracema (2019); Maassen, Eva F. (2018); Camden, Chantal (2019); Banfield, Michelle A. (2014); Zitko, Pedro (2017) |
| **Group decision-making** | Delphi method ; Nominal group technique; Consensus decision-making | Ghisoni, Marjorie (2017); Furyk, Jeremy (2018); Forsman, Anna K. (2015); Lee, C. (2019); Tuffrey-Wijne, I. (2016); van der Feltz-Cornelis, C.M. (2018); Lim, Ai Keow (2019); Elfeddali, Iman (2014); Furth, Eric F. van (2016); Wykes, Til (2015); Blum, Nathan J. (2012); Hitch, Danielle (2015); Shattuck, Paul T. (2018); Bell, Sigall K. (2018); Vasa, Roma A. (2018); Turner, Grace M. (2018); Leroi, Iracema (2019); Hart, Laura M. (2019); Pickett, James (2018) |
| **Interview and consulting** | consultation; interview; online survey; ssurvey; submitted comments | Davison, Karen M. (2017); Clavisi, Ornella (2013); Kühne, F. (2019); Banfield, Michelle A. (2011); van der Feltz-Cornelis, C.M. (2018); Lim, Ai Keow (2019); Gregório, Guilherme (2012); Paul, C. (2017); Kramer, Jessica M. (2019); Gaynes, Bradley N. (2014); Kelly, Sarah (2015); Obeid, Nicole (2020); Shattuck, Paul T. (2018); Pellicano, Elizabeth (2014); Fiorillo, Andrea (2013); Clark, Kristen D. (2019); Shah, Hiral (2016); van der Feltz-Cornelis, Christina M (2014); Leroi, Iracema (2019); Maassen, Eva F. (2018); Camden, Chantal (2019); Russell, Ginny (2018); Banfield, Michelle A. (2018); Law, Emma (2013); Banfield, Michelle A. (2014); Zitko, Pedro (2017) |
| **Literature review** | literature review | Clavisi, Ornella (2013); Kühne, F. (2019); Wykes, Til (2015); Gaynes, Bradley N. (2014); Kelly, Sarah (2015); Obeid, Nicole (2020); Shattuck, Paul T. (2018); Glandon, Douglas (2018); Reifels, Lennart (2018); Miah, Jahanara (2019); Zitko, Pedro (2017) |
| **Meetings and workshops** | workshop; meeting; thematic rountable discussions; storytelling sessions; webinars | Davison, Karen M. (2017); Ghisoni, Marjorie (2017); Clavisi, Ornella (2013); Forsman, Anna K. (2015); Lee, C. (2019); Tuffrey-Wijne, I. (2016); Mei, Cristina (2020); Crocker, Anne G. (2015); van der Feltz-Cornelis, C.M. (2018); Kramer, Jessica M. (2019); Obeid, Nicole (2020); Brophy, Lisa (2018); Jacobson, Stella (2016); Khandelwal, Sudhir (2010); Shattuck, Paul T. (2018); Glandon, Douglas (2018); Turner, Grace M. (2018); van der Feltz-Cornelis, Christina M (2014) |
| **Mixed methods research** | mixed methods design | MacGabhann, Liam (2018); Maassen, Eva F. (2018) |
| **Priority Setting Partnership** | collaborative data analysis; patient and public involvement; Canadian Dementia Priority Setting Partnership; James Lind Alliance; Participatory Action Research; Patient-centered outcomes research | Bethell, Jennifer (2018); Lim, Ai Keow (2019); Furth, Eric F. van (2016); MacGabhann, Liam (2018); Kelly, Sarah (2015); Obeid, Nicole (2020); Jacobson, Stella (2016); Hollis, Chris (2018); Miah, Jahanara (2019); Russell, Ginny (2018) |
| **Theory of change methodology** | Theory of change methodology | Leroi, Iracema (2019) |

### Table GGG. Priorities grouped by subject class

| | #priorities | # papers | papers |
|---|---|---|---|
| classification | 7 | 6 | Banfield, Michelle A. (2018); Bell, Sigall K. (2018); Fiorillo, Andrea (2013); Forsman, Anna K. (2015); Shattuck, Paul T. (2018); Tuffrey-Wijne, I. (2016) |
| complications | 21 | 16 | Banfield, Michelle A. (2014); Banfield, Michelle A. (2018); Baulac, Michel (2015); Blum, Nathan J. (2012); Camden, Chantal (2019); Clark, Kristen D. (2019); Clavisi, Ornella (2013); Elfeddali, Iman (2014); Fiorillo, Andrea (2013); Hart, Laura M. (2019); Kramer, Jessica M. (2019); Obeid, Nicole (2020); Pellicano, Elizabeth (2014); Turner, Grace M. (2018); van der Feltz-Cornelis, C.M. (2018); Vasa, Roma A. (2018) |
| diagnosis | 37 (1) | 20 | Banfield, Michelle A. (2014); Baulac, Michel (2015); Blum, Nathan J. (2012); Camden, Chantal (2019); Crocker, Anne G. (2015); Elfeddali, Iman (2014); Fiorillo, Andrea (2013); Gaynes, Bradley N. (2014); Hart, Laura M. (2019); Kelly, Sarah (2015); Law, Emma (2013); Lim, Ai Keow (2019); Maassen, Eva F. (2018); Mei, Cristina (2020); Pickett, James (2018); Shah, Hiral (2016); Tomlinson, Mark (2014); Turner, Grace M. (2018); van der Feltz-Cornelis, C.M. (2018); Vasa, Roma A. (2018) |
| diet therapy | 9 | 2 | Davison, Karen M. (2017); Kelly, Sarah (2015) |
| drug therapy | 32 | 13 | Banfield, Michelle A. (2014); Banfield, Michelle A. (2011); Banfield, Michelle A. (2018); Blum, Nathan J. (2012); Elfeddali, Iman (2014); Fiorillo, Andrea (2013); Furyk, Jeremy (2018); Jacobson, Stella (2016); Kelly, Sarah (2015); Kramer, Jessica M. (2019); Law, Emma (2013); Maassen, Eva F. (2018); van der Feltz-Cornelis, Christina M (2014) |
| economics | 16 | 13 | Banfield, Michelle A. (2011); Banfield, Michelle A. (2014); Banfield, Michelle A. (2018); Baulac, Michel (2015); Blum, Nathan J. (2012); Elfeddali, Iman (2014); Fiorillo, Andrea (2013); Ghisoni, Marjorie (2017); Gregório, Guilherme (2012); Khandelwal, Sudhir (2010); Tuffrey-Wijne, I. (2016); van der Feltz-Cornelis, Christina M (2014); Wykes, Til (2015) |
| education | 46 | 25 | Banfield, Michelle A. (2014); Banfield, Michelle A. (2011); Banfield, Michelle A. (2018); Baulac, Michel (2015); Bell, Sigall K. (2018); Bethell, Jennifer (2018); Blum, Nathan J. (2012); Camden, Chantal (2019); Clavisi, Ornella (2013); Crocker, Anne G. (2015); Furyk, Jeremy (2018); Ghisoni, Marjorie (2017); Hollis, Chris (2018); Jacobson, Stella (2016); Khandelwal, Sudhir (2010); Lee, C. (2019); Leroi, Iracema (2019); Lim, Ai Keow (2019); Maassen, Eva F. (2018); Obeid, Nicole (2020); Pellicano, Elizabeth (2014); Pickett, James (2018); Tuffrey-Wijne, I. (2016); Turner, Grace M. (2018); Wykes, Til (2015) |

| | | | |
|---|---|---|---|
| epidemiology | 8 | 6 | Bell, Sigall K. (2018); Fiorillo, Andrea (2013); Gregório, Guilherme (2012); Khandelwal, Sudhir (2010); Pickett, James (2018); Reifels, Lennart (2018) |
| ethnology | 9 | 8 | Banfield, Michelle A. (2018); Brophy, Lisa (2018); Fiorillo, Andrea (2013); Hart, Laura M. (2019); Hitch, Danielle (2015); Mei, Cristina (2020); Shah, Hiral (2016); Zitko, Pedro (2017) |
| etiology | 19 | 15 | Banfield, Michelle A. (2014); Banfield, Michelle A. (2018); Baulac, Michel (2015); Blum, Nathan J. (2012); Clark, Kristen D. (2019); Fiorillo, Andrea (2013); Furyk, Jeremy (2018); Hart, Laura M. (2019); Maassen, Eva F. (2018); Reifels, Lennart (2018); Shah, Hiral (2016); Tomlinson, Mark (2014); van der Feltz-Cornelis, Christina M (2014); Vasa, Roma A. (2018); Zitko, Pedro (2017) |
| genetics | 4 | 3 | Baulac, Michel (2015); Blum, Nathan J. (2012); Fiorillo, Andrea (2013) |
| legislation and jurisprudence | 6 | 3 | Banfield, Michelle A. (2018); Baulac, Michel (2015); Brophy, Lisa (2018) |
| methods | 65 | 30 | Banfield, Michelle A. (2014); Banfield, Michelle A. (2018); Baulac, Michel (2015); Bethell, Jennifer (2018); Blum, Nathan J. (2012); Camden, Chantal (2019); Elfeddali, Iman (2014); Fiorillo, Andrea (2013); Forsman, Anna K. (2015); Gaynes, Bradley N. (2014); Ghisoni, Marjorie (2017); Glandon, Douglas (2018); Gregório, Guilherme (2012); Hitch, Danielle (2015); Hollis, Chris (2018); Khandelwal, Sudhir (2010); Kramer, Jessica M. (2019); Lee, C. (2019); Lim, Ai Keow (2019); Maassen, Eva F. (2018); Pickett, James (2018); Shah, Hiral (2016); Tol, Wietse A. (2012); Tomlinson, Mark (2014); Tuffrey-Wijne, I. (2016); van der Feltz-Cornelis, C.M. (2018); van der Feltz-Cornelis, Christina M (2014); Vasa, Roma A. (2018); Wykes, Til (2015); Zitko, Pedro (2017) |
| organization and administration | 20 | 9 | Banfield, Michelle A. (2018); Baulac, Michel (2015); Bethell, Jennifer (2018); Crocker, Anne G. (2015); Ghisoni, Marjorie (2017); Glandon, Douglas (2018); Gregório, Guilherme (2012); Leroi, Iracema (2019); Shattuck, Paul T. (2018) |
| physiopathology | 9 | 7 | Baulac, Michel (2015); Camden, Chantal (2019); Elfeddali, Iman (2014); Furth, Eric F. van (2016); Mei, Cristina (2020); Shah, Hiral (2016); Wykes, Til (2015) |
| prevention and control | 34 | 23 | Banfield, Michelle A. (2011); Banfield, Michelle A. (2018); Blum, Nathan J. (2012); Camden, Chantal (2019); Crocker, Anne G. (2015); Davison, Karen M. (2017); Elfeddali, Iman (2014); Fiorillo, Andrea (2013); Furth, Eric F. van (2016); Ghisoni, Marjorie (2017); Gregório, Guilherme (2012); Hart, Laura M. (2019); Lee, C. (2019); Leroi, Iracema (2019); Lim, Ai Keow (2019); Mei, Cristina (2020); Obeid, Nicole (2020); Reifels, Lennart (2018); Shah, Hiral (2016); Tol, Wietse A. (2012); Tomlinson, Mark (2014); Wykes, Til (2015); Zitko, Pedro (2017) |

| | | | |
|---|---|---|---|
| psychology | 50 | 26 | Banfield, Michelle A. (2014); Banfield, Michelle A. (2018); Bell, Sigall K. (2018); Bethell, Jennifer (2018); Blum, Nathan J. (2012); Camden, Chantal (2019); Clark, Kristen D. (2019); Clavisi, Ornella (2013); Elfeddali, Iman (2014); Fiorillo, Andrea (2013); Ghisoni, Marjorie (2017); Glandon, Douglas (2018); Hart, Laura M. (2019); Kramer, Jessica M. (2019); Kühne, F. (2019); Law, Emma (2013); Lee, C. (2019); Lim, Ai Keow (2019); Maassen, Eva F. (2018); Pellicano, Elizabeth (2014); Pickett, James (2018); Reifels, Lennart (2018); Shah, Hiral (2016); Tol, Wietse A. (2012); Turner, Grace M. (2018); Zitko, Pedro (2017) |
| rehabilitation | 44 | 23 | Banfield, Michelle A. (2011); Banfield, Michelle A. (2018); Blum, Nathan J. (2012); Camden, Chantal (2019); Clavisi, Ornella (2013); Crocker, Anne G. (2015); Elfeddali, Iman (2014); Fiorillo, Andrea (2013); Furth, Eric F. van (2016); Ghisoni, Marjorie (2017); Hitch, Danielle (2015); Hollis, Chris (2018); Jacobson, Stella (2016); Kelly, Sarah (2015); Khandelwal, Sudhir (2010); Kramer, Jessica M. (2019); Lee, C. (2019); Maassen, Eva F. (2018); Pellicano, Elizabeth (2014); Shah, Hiral (2016); Shattuck, Paul T. (2018); Turner, Grace M. (2018); Vasa, Roma A. (2018) |
| standards | 70 | 29 | Banfield, Michelle A. (2011); Banfield, Michelle A. (2018); Bethell, Jennifer (2018); Blum, Nathan J. (2012); Brophy, Lisa (2018); Camden, Chantal (2019); Crocker, Anne G. (2015); Elfeddali, Iman (2014); Fiorillo, Andrea (2013); Forsman, Anna K. (2015); Furth, Eric F. van (2016); Gaynes, Bradley N. (2014); Glandon, Douglas (2018); Kelly, Sarah (2015); Kramer, Jessica M. (2019); Lee, C. (2019); Lim, Ai Keow (2019); Maassen, Eva F. (2018); Obeid, Nicole (2020); Pickett, James (2018); Reifels, Lennart (2018); Shah, Hiral (2016); Shattuck, Paul T. (2018); Tol, Wietse A. (2012); Tuffrey-Wijne, I. (2016); Turner, Grace M. (2018); van der Feltz-Cornelis, Christina M (2014); Vasa, Roma A. (2018); Zitko, Pedro (2017) |
| statistics and numerical data | 2 | 2 | Obeid, Nicole (2020); Pickett, James (2018) |
| supply and distribution | 41 | 23 | Banfield, Michelle A. (2014); Banfield, Michelle A. (2018); Baulac, Michel (2015); Bethell, Jennifer (2018); Camden, Chantal (2019); Clavisi, Ornella (2013); Crocker, Anne G. (2015); Elfeddali, Iman (2014); Ghisoni, Marjorie (2017); Hollis, Chris (2018); Khandelwal, Sudhir (2010); Kramer, Jessica M. (2019); Lee, C. (2019); Leroi, Iracema (2019); Mei, Cristina (2020); Obeid, Nicole (2020); Pellicano, Elizabeth (2014); Pickett, James (2018); Shattuck, Paul T. (2018); Tomlinson, Mark (2014); Tuffrey-Wijne, I. (2016); Turner, Grace M. (2018); Zitko, Pedro (2017) |

| | | | |
|---|---|---|---|
| therapy | 105 | 31 | Banfield, Michelle A. (2014); Banfield, Michelle A. (2011); Banfield, Michelle A. (2018); Baulac, Michel (2015); Bethell, Jennifer (2018); Blum, Nathan J. (2012); Brophy, Lisa (2018); Camden, Chantal (2019); Clavisi, Ornella (2013); Elfeddali, Iman (2014); Fiorillo, Andrea (2013); Furth, Eric F. van (2016); Gaynes, Bradley N. (2014); Gregório, Guilherme (2012); Hart, Laura M. (2019); Hitch, Danielle (2015); Hollis, Chris (2018); Jacobson, Stella (2016); Kelly, Sarah (2015); Kramer, Jessica M. (2019); Kühne, F. (2019); Maassen, Eva F. (2018); Mei, Cristina (2020); Obeid, Nicole (2020); Pellicano, Elizabeth (2014); Pickett, James (2018); Shah, Hiral (2016); Turner, Grace M. (2018); van der Feltz-Cornelis, C.M. (2018); van der Feltz-Cornelis, Christina M (2014); Vasa, Roma A. (2018) |
| other | 14 | 8 | Banfield, Michelle A. (2018); Elfeddali, Iman (2014); Fiorillo, Andrea (2013); Hitch, Danielle (2015); Lee, C. (2019); Maassen, Eva F. (2018); Mei, Cristina (2020); Tuffrey-Wijne, I. (2016) |

#### Table BBB. MeSH subheadings used in classifying priorities

| subheading | definition |
|---|---|
| **classification** | Used for taxonomic or other systematic or hierarchical classification systems. |
| **complications** | Used with diseases to indicate conditions that co-exist or follow, i.e., co-existing diseases, complications, or sequelae. |
| **diagnosis** | Used with diseases for all aspects of diagnosis, including examination, differential diagnosis and prognosis. Excludes diagnosis using imaging techniques (e.g. radiography, scintigraphy, and ultrasonography) for which diagnostic imaging is used. |
| **diagnostic imaging** | Used for the visualization of an anatomical structure or for the diagnosis of disease. Commonly used imaging techniques include radiography, radionuclide imaging, thermography, tomography, and ultrasonography. |
| **drug therapy** | Used with disease headings for the treatment of disease by the administration of drugs, chemicals, and antibiotics. For diet therapy and radiotherapy, use specific subheadings. Excludes immunotherapy for which therapy is used. |
| **economics** | Used for the economic aspects of any subject, as well as for all aspects of financial management. It includes the raising or providing of funds. |
| **education** | Used for education, training programs, and courses in various fields and disciplines, and for training groups of persons. |
| **epidemiology** | Used with human and veterinary diseases for the distribution of disease, factors which cause disease, and the attributes of disease in defined populations; includes incidence, frequency, prevalence, endemic and epidemic outbreaks; also surveys and estimates of morbidity in geographic areas and in specified populations. Used also with geographical headings for the location of epidemiologic aspects of a disease. Excludes mortality for which mortality is used. |
| **ethnology** | Used with diseases for ethnic, cultural, or anthropological aspects, and with geographic headings to indicate the place of origin of a group of people. |
| **etiology** | Used with diseases for causative agents including microorganisms and includes environmental and social factors and personal habits as contributing factors. It includes pathogenesis. |
| **genetics** | Used for mechanisms of heredity and the genetics of organisms, for the genetic basis of normal and pathologic states, and for the genetic aspects of endogenous chemicals. It includes biochemical and molecular influence on genetic material. |
| **legislation and jurisprudence** | Used for laws, statutes, ordinances, or government regulations, as well as for legal controversy and court decisions. |
| **methods** | Used with techniques, procedures, and programs for methods. |
| **organization and administration** | Used for administrative structure and management. |
| **physiopathology** | Used with organs and diseases for disordered function in disease states. |

| subheading | definition |
|---|---|
| **prevention and control** | Used with disease headings for increasing human or animal resistance against disease (e.g., immunization), for control of transmission agents, for prevention and control of environmental hazards, or for prevention and control of social factors leading to disease. It includes preventive measures in individual cases. |
| **psychology** | Used with non-psychiatric diseases, techniques, and named groups for psychologic, psychiatric, psychosomatic, psychosocial, behavioral, and emotional aspects, and with psychiatric disease for psychologic aspects; used also with animal terms for animal behavior and psychology. |
| **rehabilitation** | Used with diseases and surgical procedures for restoration of function of the individual. |
| **standards** | Used with facilities, personnel, and program headings for the development, testing, and application of standards of adequacy or acceptable performance and with chemicals and drugs for standards of identification, quality, and potency. It includes health or safety standards in industries and occupations. |
| **statistics and numerical data** | Used with non-disease headings for the expression of numerical values that describe particular sets or groups of data. It includes level of use of equipment and supplies, facilities and services and procedures and techniques. It excludes supply or demand for which supply and distribution is used |
| **supply and distribution** | Used for the quantitative availability and distribution of material, equipment, health services, personnel, and facilities. It excludes food supply and water supply in industries and occupations. |
| **therapy** | Used with diseases for therapeutic interventions except drug therapy, diet therapy, radiotherapy, and surgery, for which specific subheadings exist. The concept is also used for articles and books dealing with multiple therapies. |
